# The Evolving Role of Telehealth in Routine Health Care Delivery Among US Adults, 2022–2024

**DOI:** 10.64898/2026.11.09.26367679

**Authors:** Shengyang Wang, Wenxue Lin, Lin Ge

**Author notes:** **Corresponding Author:** Lin Ge, Ph.D., Department of Epidemiology and Biostatistics Indiana University School of Public Health 1025 E 7th St, Bloomington, IN 47405.

## Abstract

**Background:** Telehealth use expanded rapidly during the COVID-19 pandemic, but its role following the end of the public health emergency (PHE) remains incompletely understood. This study examined changes in telehealth utilization among US adults between 2022 and 2024, characteristics associated with use, and changes in self-reported reasons for using telehealth to understand its evolving role.

**Methods:** This serial cross-sectional study analyzed nationally representative data from 13 141 US adults participating in the Health Information National Trends Survey (HINTS) in 2022, during the COVID-19 PHE, and 2024, after the PHE ended. Survey-weighted analyses were used to estimate telehealth utilization and self-reported reasons for use. Survey-weighted multivariable logistic regression was used to examine changes between survey cycles and sociodemographic and health-related characteristics associated with telehealth utilization. Multiple imputation was used to address missing data.

**Results:** Telehealth utilization declined from 39.3% in 2022 to 34.5% in 2024; after adjustment for sociodemographic and health-related characteristics, the odds of telehealth use were significantly lower in 2024 than in 2022 (adjusted odds ratio [aOR], 0.81; 95% CI, 0.69-0.95). Telehealth utilization differed across population groups. Higher odds of use were observed among adults aged 35 to 49 years (aOR, 1.47; 95% CI, 1.19-1.82), those with a college degree or higher (aOR, 1.93; 95% CI, 1.31-2.86), those with diagnosed depression or anxiety (aOR, 2.86; 95% CI, 2.46-3.33), those with a history of cancer (aOR, 1.40; 95% CI, 1.15-1.69), and insured adults (aOR, 1.52; 95% CI, 1.13-2.04). All self-reported reasons for telehealth use declined between 2022 and 2024. Infection avoidance decreased from 50.2% to 19.1% and provider recommendation or requirement from 72.8% to 47.2%, whereas convenience showed a smaller decline from 65.6% to 56.2% and became the leading reported reason for telehealth use in 2024.

**Conclusions:** Telehealth utilization declined between 2022 and 2024, while the relative persistence of convenience as a reason for use suggests a shift away from pandemic-driven demand toward more selective integration into routine health care. Differences in utilization across population groups suggest that telehealth may address some barriers to conventional care while access remains uneven. These findings highlight the evolving role of telehealth after the COVID-19 PHE and the importance of supporting its appropriate and equitable integration into routine care.

## Introduction

The use of digital technologies in health care has expanded rapidly over the past decade^1,2^. Innovations such as wearable devices, mobile health applications, and real-time audiovisual communication platforms have become increasingly integrated into routine health care and daily life^3–6^. Among these innovations, telehealth has emerged as an approach to delivering health care remotely^7,8^, with the potential to improve access, convenience, and flexibility by reducing geographic and time-related barriers^9–11^. Despite these potential advantages, telehealth adoption in the United States remained limited before the COVID-19 pandemic^12^. Among enrollees in large employer health plans, the proportion using at least 1 telehealth service increased from only 0.8% in 2016 to 2.4% in 2018^13^. However, the COVID-19 pandemic marked a dramatic turning point in telehealth use. Federal policy changes expanded reimbursement and removed geographic and regulatory barriers, including allowing Medicare beneficiaries to receive telehealth services from home, while regulatory flexibilities facilitated the use of remote communication technologies for clinical care^14–17^. Against this rapidly changing policy and health care environment, Medicare telehealth utilization increased from approximately 840 000 visits in 2019 to 52.7 million visits in 2020, representing an approximately 63-fold increase^18^. During the COVID-19 public health emergency (PHE), telehealth became an important mechanism for maintaining continuity of care while reducing potential infection exposure and disruptions to in-person health care^19^.

Following the end of the COVID-19 PHE, the conditions that had supported rapid telehealth expansion changed substantially^12,20,21^. Infection-related concerns diminished, routine in-person care resumed, and pandemic-era telehealth policies were modified or discontinued^22^. Recent studies have documented declining telehealth utilization and differences in use across population groups^23–25^. These changes have also raised broader questions about telehealth’s role beyond the pandemic. After its rapid expansion during the PHE, whether declining utilization indicates a diminishing role for telehealth or reflects a transition toward a different role within routine health care delivery remains unclear. Moreover, although telehealth can address some limitations of conventional in-person care, including geographic and time-related barriers, its increasing integration into health care may introduce or reinforce distinct barriers to equitable access^26^. Existing studies have characterized individual aspects of post-pandemic telehealth use, including utilization trends and differences across population groups, but these dimensions have often been examined separately^25,27^. Consequently, how these patterns collectively characterize telehealth’s role as health care delivery returns to more routine conditions, and whether they reveal persistent or emerging challenges to equitable access, remains incompletely understood.

Characterizing this transition requires more than examining changes in utilization or characteristics associated with use in isolation. It also requires understanding the reasons patients report for using telehealth, what these reasons may reveal about its role in health care delivery, and how patterns of telehealth use differ across population groups during and after the PHE. To provide an integrated assessment of these dimensions, we analyzed nationally representative Health Information National Trends Survey (HINTS) data collected in 2022, during the COVID-19 PHE, and 2024, after the PHE ended. The nationally representative HINTS data allowed these dimensions of telehealth use to be examined together among US adults. We examined changes in overall telehealth utilization between the 2 periods, sociodemographic and health-related characteristics associated with use, changes in self-reported reasons for using telehealth, and patterns of telehealth utilization across population subgroups in 2022 and 2024. By considering these findings together and in the context of existing evidence, this study aimed to characterize telehealth’s evolving role in routine health care delivery and its implications for health care access and equity beyond the PHE.

## Methods

### Data Source and Study Population

This study analyzed data from the Health Information National Trends Survey (HINTS), a nationally representative cross-sectional survey of non-institutionalized US adults aged 18 years or older^28^. Details of the survey methodology have been described previously^29^. We used data from HINTS 6 (2022) and HINTS 7 (2024), which included 6 252 and 7 278 respondents and had response rates of 28.1% and 27.3%, respectively^30,31^. Telehealth utilization data were available for 6 046 respondents in HINTS 6 and 7 095 respondents in HINTS 7. Following HINTS analytic guidance, data from the 2 survey cycles were combined, yielding a primary analytic sample of 13 141 participants. This study followed the Strengthening the Reporting of Observational Studies in Epidemiology (STROBE) reporting guideline for cross-sectional studies^32^.

### Study Variables and Outcomes

The primary outcome was telehealth utilization, assessed using the survey question, “In the past 12 months, did you receive care from a doctor or health professional using telehealth?” Respondents who reported receiving care by video, telephone, or both were classified as telehealth users; those who reported “No” were classified as non-users.

Secondary outcomes were self-reported reasons for telehealth use among telehealth users. Participants were asked whether they used telehealth because (1) a health care provider recommended or required it, (2) they wanted advice regarding the need for in-person medical care, (3) they wanted to avoid potential infection exposure, (4) telehealth was more convenient than an in-person visit, or (5) family members or caregivers could participate in the appointment.

Covariates included sociodemographic and health-related characteristics: age (18-34, 35-49, 50-64, 65-74, or ≥75 years), sex (female or male), race and ethnicity (Hispanic, non-Hispanic Black, non-Hispanic White, or other), educational attainment (less than high school, high school graduate, some college, or college graduate or higher), annual household income (<$20 000, $20 000 to <$35 000, $35 000 to <$50 000, $50 000 to <$75 000, or ≥$75 000), and Census region (Northeast, Midwest, South, or West). Additional covariates included health insurance status (insured or uninsured), history of cancer (yes or no), diagnosis of depression or anxiety (yes or no), and weekly work hours (≤35 or >35 hours). Health-related covariates were selected based on factors commonly examined in previous studies of telehealth utilization^33^.

### Missing Data and Multiple Imputation

Missingness across study variables ranged from 1.25% to 12.35%. Consistent with HINTS guidance, missing data were assumed to be missing at random (MAR)^34^. Multiple Imputation by Chained Equations (MICE) was used to address missingness and preserve statistical power. The imputation model included all covariates, telehealth utilization and survey cycles; fully observed variables and outcomes were included as predictors but were not imputed. Binary variables were imputed using classic logistic regression, multinomial variables using polytomous logistic regression, and variables with sparse categories using classification and regression tree (CART) models. Ten imputed datasets were generated, analyzed separately, and pooled using Rubin’s rules. Multiple imputation was performed using the *MICE* package in R, version 4.5.3.

### Statistical Analysis

All analyses incorporated the combined full-sample weights and jackknife replicate weights to account for the complex survey design and generate nationally representative estimates of US adults. Full-sample weights were used to produce population estimates, whereas jackknife replicate weights were used for variance estimation.

Descriptive analyses were conducted separately for HINTS 6 and HINTS 7 to summarize participant characteristics according to telehealth utilization status. Unweighted counts and survey-weighted percentages were calculated, and Rao–Scott χ^2^ tests were used to assess associations between participant characteristics and telehealth utilization within each survey cycle.

The primary analysis used survey-weighted logistic regression to examine changes in telehealth utilization between survey cycles and factors associated with its use. The model included telehealth utilization as the outcome and survey cycle (HINTS 7 vs HINTS 6), sociodemographic characteristics and health-related factors as covariates.

Secondary analyses were restricted to 5 031 telehealth users (HINTS 6: n = 2 517; HINTS 7: n = 2 514). Survey-weighted percentages were calculated for each self-reported reason for telehealth use. Separate survey-weighted logistic regression models were fitted for each reason to estimate changes between survey cycles after adjustment for the covariates included in the primary model.

All regression models were fitted separately in each imputed dataset, and regression coefficients and standard errors were combined using Rubin’s rules. Statistical significance was defined as a 2-sided P < .05. All analyses were performed using R, version 4.5.3.

## Results

### Characteristics and Temporal Changes in Telehealth Utilization

Table 1 presents participant characteristics according to telehealth utilization status in 2022 and 2024. Overall, similar patterns were observed across both survey cycles. Telehealth use was more common among women than men and among participants with higher educational attainment. Participants aged 35 to 49 years represented the largest proportion of telehealth users, whereas utilization was lower among older adults. Diagnosed depression or anxiety was reported more prevalent among telehealth users than non-users. In contrast, work hours showed a reversed pattern between 2022 and 2024. Cancer history, and health insurance status showed similar patterns across the two survey years.

**Table 1.** Participant Characteristics by Telehealth Utilization Status in 2022 and 2024.

|  | 2022 (HINTS 6)<sup>a</sup>, N = 6046 |  |  |  | 2024 (HINTS 7)<sup>a</sup>, N = 7095 |  |  |  |
|---|---|---|---|---|---|---|---|---|
| Characteristic | Non-users, No. (weighted %) | Users, No. (weighted %) | P value<sup>b</sup> | Missing, No.(weighted %)<sup>c</sup> | Non-users, No. (weighted %) | Users, No. (weighted %) | P value<sup>b</sup> | Missing, No.(weighted %)<sup>c</sup> |
| Overall | 3529 (60.7) | 2517 (39.3) | NA | NA | 4581 (65.5) | 2514 (34.5) | NA | NA |
| Sex |  |  | <.001 | 410 (6.6) |  |  | <.001 | 615 (8.5) |
| Female | 1919 (46.4) | 1546 (56.9) |  |  | 2464 (45.5) | 1515 (54.9) |  |  |
| Male | 1446 (53.6) | 832 (43.1) |  |  | 1811 (54.5) | 819 (45.1) |  |  |
| Education |  |  | <.001 | 404 (6.5) |  |  | <.001 | 583 (8.0) |
| <High school | 253 (7.7) | 116 (4.9) |  |  | 297 (6.9) | 137 (4.7) |  |  |
| High school | 691 (23.4) | 351 (18.9) |  |  | 808 (23.4) | 298 (17.4) |  |  |
| Some college | 980 (39.4) | 662 (38.0) |  |  | 1255 (38.5) | 669 (38.8) |  |  |
| College degree or higher | 1449 (29.5) | 1248 (38.2) |  |  | 1934 (31.2) | 1243 (39.1) |  |  |
| Age |  |  | .035 | 98 (1.6) |  |  | <.001 | 613 (8.4) |
| 18–34 | 523 (27.1) | 376 (23.3) |  |  | 657 (26.0) | 421 (24.1) |  |  |
| 35–49 | 649 (22.8) | 555 (28.9) |  |  | 779 (23.8) | 575 (31.1) |  |  |
| 50–64 | 1009 (27.8) | 718 (27.4) |  |  | 1098 (26.3) | 615 (26.5) |  |  |
| 65–74 | 788 (13.4) | 527 (12.6) |  |  | 991 (13.6) | 458 (11.8) |  |  |
| ≥75 | 513 (8.9) | 306 (7.9) |  |  | 749 (10.3) | 269 (6.6) |  |  |
| Household income, \$ |  |  | .042 | 732 (11.7) |  |  | .157 | 899 (12.3) |
| <20 000 | 555 (14.7) | 368 (13.2) |  |  | 720 (15.1) | 388 (15.0) |  |  |
| 20 000–34 999 | 461 (11.6) | 254 (10.0) |  |  | 557 (12.9) | 239 (9.1) |  |  |
| 35 000–49 999 | 434 (12.0) | 290 (11.7) |  |  | 522 (11.2) | 254 (12.3) |  |  |
| 50 000–74 999 | 562 (19.9) | 364 (15.9) |  |  | 651 (15.5) | 373 (15.8) |  |  |
| ≥75 000 | 1161 (41.9) | 982 (49.2) |  |  | 1614 (45.3) | 1017 (47.8) |  |  |
| Race and ethnicity |  |  | .450 | 687 (11.0) |  |  | .381 | 840 (11.5) |
| Hispanic | 542 (16.8) | 439 (17.1) |  |  | 808 (18.0) | 504 (16.7) |  |  |
| Non-Hispanic White | 1853 (61.4) | 1307 (61.7) |  |  | 2336 (61.2) | 1188 (59.3) |  |  |
| Non-Hispanic Black | 529 (11.7) | 344 (9.7) |  |  | 595 (10.2) | 367 (12.5) |  |  |
| Other<sup>d</sup> | 274 (10.1) | 190 (11.5) |  |  | 366 (10.5) | 231 (11.5) |  |  |
| Census region |  |  | <.001 | 0 (0.0) |  |  | .193 | 0 (0.0) |
| Northeast | 507 (16.2) | 394 (19.3) |  |  | 656 (17.7) | 375 (16.6) |  |  |
| Midwest | 691 (24.6) | 336 (15.5) |  |  | 823 (21.3) | 372 (19.4) |  |  |
| South | 1647 (38.9) | 1087 (36.4) |  |  | 2166 (39.1) | 1079 (38.0) |  |  |
| West | 684 (20.3) | 700 (28.9) |  |  | 936 (21.9) | 688 (26.1) |  |  |
| Depression or anxiety diagnosis |  |  | <.001 | 241 (3.9) |  |  | <.001 | 381 (5.2) |
| No | 2798 (80.7) | 1558 (60.8) |  |  | 3511 (78.2) | 1482 (57.6) |  |  |
| Yes | 658 (19.3) | 898 (39.2) |  |  | 908 (21.8) | 941 (42.4) |  |  |
| Cancer history |  |  | .007 | 370 (5.9) |  |  | .063 | 532 (7.3) |
| No | 2899 (91.2) | 2005 (87.7) |  |  | 3668 (90.9) | 1966 (89.2) |  |  |
| Yes | 487 (8.8) | 392 (12.3) |  |  | 664 (9.1) | 396 (10.8) |  |  |
| Health insurance |  |  | <.001 | 126 (2.0) |  |  | .951 | 91 (1.3) |
| No | 360 (13.8) | 143 (5.7) |  |  | 439 (8.6) | 213 (8.5) |  |  |
| Yes | 3147 (86.2) | 2361 (94.3) |  |  | 4120 (91.4) | 2286 (91.5) |  |  |
| Weekly work hours |  |  | .081 | 412 (6.6) |  |  | .854 | 0 (0.0) |
| ≤35 | 1740 (43.6) | 1250 (47.9) |  |  | 2771 (51.2) | 1406 (50.8) |  |  |
| >35 | 1621 (56.4) | 1129 (52.1) |  |  | 1810 (48.8) | 1108 (49.2) |  |  |

<sup>a</sup> Data were obtained from the Health Information National Trends Survey HINTS 6, conducted in 2022, and HINTS 7, conducted in 2024. Weighted percentages may not sum to 100% because of rounding. Telehealth utilization status was missing for 206 participants in 2022 and 183 participants in 2024.
<sup>b</sup> *P* values were calculated using survey-weighted Rao–Scott χ² tests and represent the overall association between each participant characteristic and telehealth utilization status within the corresponding survey year.
<sup>c</sup> Missing percentages represent the percentage of participants with missing information for each characteristic in the corresponding survey year.
<sup>d</sup> Other race or multiple races includes non-Hispanic respondents who identified as American Indian or Alaska Native, Asian, Native Hawaiian or other Pacific Islander, another race, or more than one race.

After adjustment for sociodemographic and health-related factors, telehealth utilization was significantly lower in 2024 than in 2022 (aOR, 0.81; 95% CI, 0.69-0.95; *P* = .008) (Figure 1).

**Figure 1.**
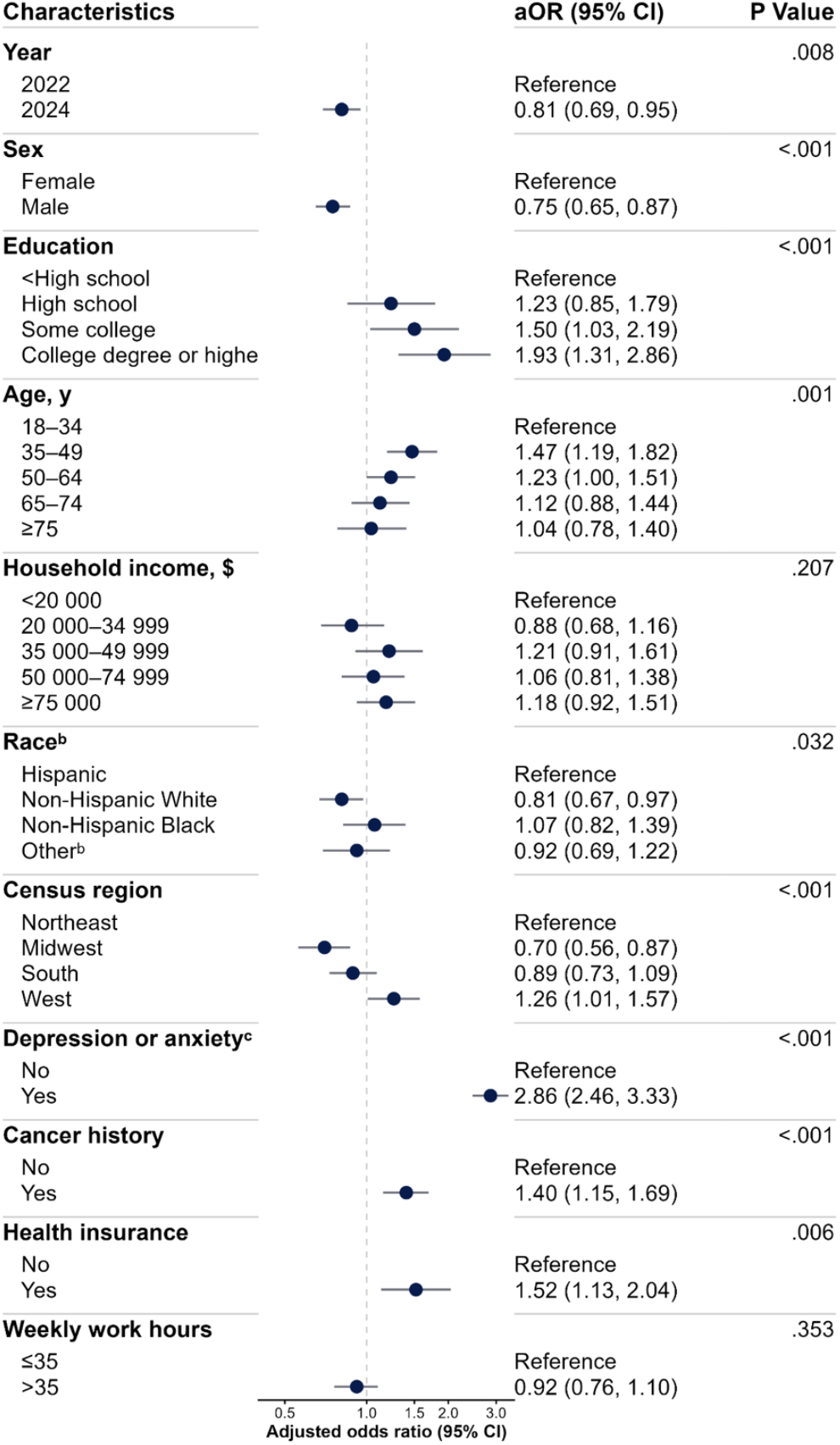
Adjusted Associations of Participant Characteristics and Survey Year with Telehealth Utilization, 2022 and 2024^a^. Adjusted odds ratios (aORs) and 95% CIs were estimated using survey-weighted logistic regression models. *P* values were obtained from survey-weighted Wald tests of the overall association between each characteristic and telehealth utilization. ^a^ Data were obtained from the Health Information National Trends Survey 6, conducted in 2022, and Health Information National Trends Survey 7, conducted in 2024. For survey year, 2022 was the reference category; the aOR for 2024 represents telehealth utilization in 2024 compared with 2022. ^b^ Race refers to race and ethnicity. Other race or multiple races includes the remaining non-Hispanic racial groups and respondents who reported multiple races. ^c^ Depression or anxiety indicates a self-reported diagnosis of depression or anxiety.

Several sociodemographic characteristics were associated with telehealth utilization. Compared with adults aged 18 to 34 years, those aged 35 to 49 years had the highest odds of telehealth use (aged 35-49 years: aOR, 1.47; 95% CI, 1.19-1.82). Men had lower odds of telehealth utilization than women (aOR, 0.75; 95% CI, 0.65-0.87; *P* <.001). The odds of telehealth use increased with educational attainment, with the highest odds observed among participants who were college graduates or had higher educational attainment compared with those who had less than a high school education (≥ College: aOR, 1.93; 95% CI, 1.31-2.86). Compared with Hispanic participants, non-Hispanic White participants had lower odds of telehealth use (Non-Hispanic White: aOR, 0.81; 95% CI, 0.67-0.97), whereas no significant differences were observed for other racial and ethnic groups. Compared with participants residing in the Northeast, those residing in the West had highest odds of telehealth use (West: aOR, 1.26; 95% CI, 1.01-1.57) (Figure 1).

Several health-related characteristics were also associated with telehealth utilization. Participants with a diagnosis of depression or anxiety had higher odds of telehealth use than those without either diagnosis (aOR, 2.86; 95% CI, 2.46-3.33; *P* < .001). Higher odds were also observed among participants with health insurance (being insured: aOR, 1.52; 95% CI, 1.13-2.04; *P* = .006) and those with a history of cancer (having cancer history: aOR, 1.40; 95% CI, 1.15-1.69; *P* <.001). In contrast, weekly work hours and household income were not significantly associated with telehealth utilization (Figure 1).

### Temporal Changes in Self-reported Reasons for Telehealth Use

In 2022, the most frequently reported reasons for telehealth use were health care provider recommendation or requirement (72.8%), convenience (65.6%), and avoidance of infection exposure (50.2%). Although all self-reported reasons had lower weighted percentages in 2024 than 2022, the magnitude of decline varied substantially. Avoidance of infection exposure showed the largest decrease, declining from 50.2% to 19.1% (31.1 percentage points), followed by health care provider recommendation or requirement, which declined from 72.8% to 47.2% (25.6 percentage points). In contrast, convenience showed the smallest decrease, declining from 65.6% to 56.2% (9.4 percentage points), and became the most frequently reported reason in 2024 (Figure 2A).

**Figure 2.**
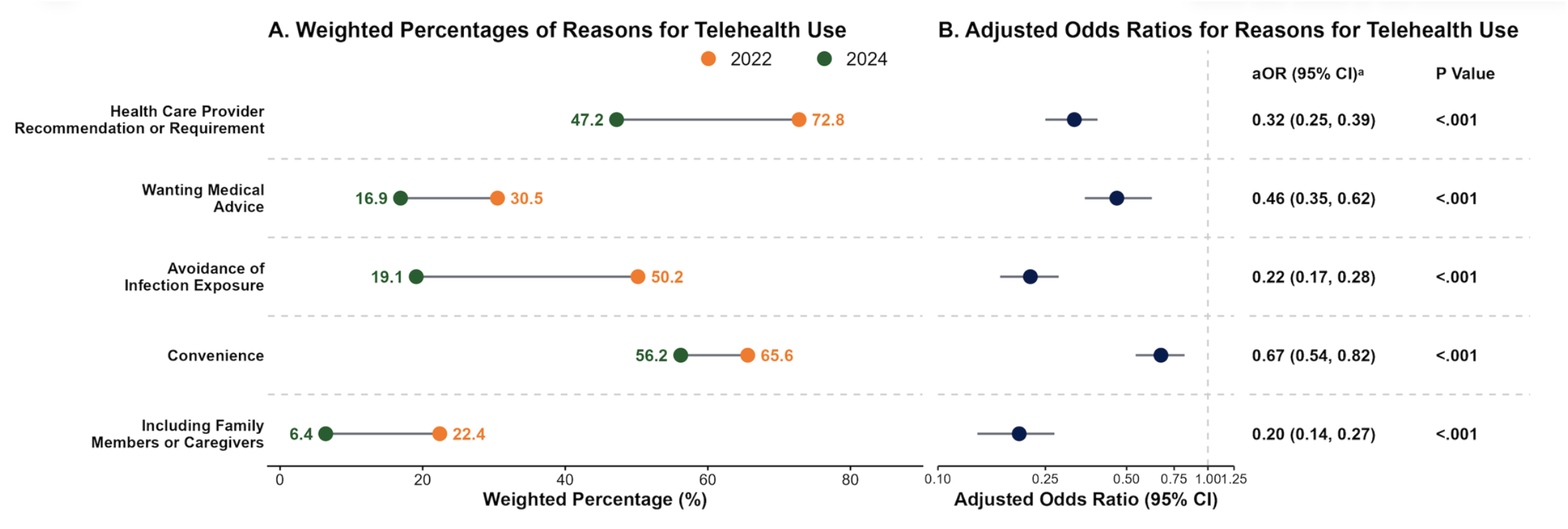
Changes in Self-reported Reasons for Telehealth Use Between 2022 and 2024.

The adjusted analyses were consistent with the descriptive findings. All self-reported reasons for telehealth use had significantly lower odds of being reported in 2024 than in 2022. Convenience exhibited the smallest decline (aOR, 0.67; 95% CI, 0.54-0.82; *P* < .001), whereas health care provider recommendation or requirement (aOR, 0.32; 95% CI, 0.25-0.39; *P* < .001) and avoidance of infection exposure (aOR, 0.22; 95% CI, 0.17-0.28; *P* < .001) had substantially lower odds of being reported in 2024 (Figure 2B; Complete adjusted model results are provided in eTables 1-5 in Supplement 1).

## Discussion

In this nationally representative study of US adults, telehealth utilization declined from 2022 to 2024, while utilization differed across population subgroups. Higher utilization among some populations may reflect ongoing clinical needs, whereas lower utilization among others may reflect differences in access, need, or preferences for telehealth. At the same time, changes in self-reported reasons for use provide insight into how the role of telehealth may be evolving. Although all reported reasons declined, convenience showed greater relative persistence and became the leading reason for telehealth use in 2024. Together, these findings suggest that telehealth may be transitioning from broad, emergency-driven adoption toward a more selective role in routine health care delivery, while differences in utilization across population groups raise important questions about equitable access.

Telehealth utilization declined from 2022 to 2024, consistent with recent studies^25,27^. This decline may reflect normalization following emergency-driven expansion rather than the disappearance of telehealth from health care delivery. During the COVID-19 PHE, expanded reimbursement, regulatory flexibility, concerns about infection, and disruptions to in-person care facilitated rapid telehealth adoption^12,14^. As the PHE ended, infection-related concerns diminished, in-person care resumed, and some temporary telehealth policies were modified or discontinued^20,35,36^. Evidence from health system and national physician surveys suggests that telehealth use, although below pandemic peaks, remains substantially above prepandemic levels^25,27,37^. A similar pattern was observed at the University of California, San Francisco, where telehealth increased from approximately 2% of outpatient visits before the pandemic to 60% during the pandemic and subsequently stabilized at approximately 25%^38^. Similarly, an American Medical Association (AMA) report showed that the proportion of physicians using telehealth in 2024 remained nearly 3 times the prepandemic level^39^. Together, these patterns suggest that declining utilization may represent a transition from emergency-driven expansion toward a more established, but less intensive, role in routine health care delivery.

Changes in self-reported reasons for telehealth use provide additional insight into this evolving role. Although all reasons were reported less frequently in 2024, provider recommendation or requirement and infection avoidance showed the largest declines, whereas convenience declined less and became the leading reported reason for telehealth use. This shift suggests that motivations for telehealth use may be moving away from pandemic-specific circumstances toward its practical advantages in health care delivery. Consistent with this interpretation, a 2024 AMA survey reported greater telehealth use in specialties such as psychiatry (85.9%), endocrinology (24.2%), gastroenterology (20.4%), and general medicine (20.1%) and lower use in anesthesia (6.2%), surgery (4.7%), and emergency medicine (4.3%)^39^. These differences may reflect the greater suitability of telehealth for services that can be delivered without physical procedures or examinations, such as consultation, medication management, and follow-up care. Telehealth is therefore unlikely to replace in-person care uniformly but may instead be selectively integrated into clinical services for which remote delivery is feasible and can reduce travel and time burdens. The emergence of convenience as the leading reported reason for use in 2024, as pandemic-related motivations became less prominent, further supports a more selective and practical role for telehealth in routine health care delivery.

Differences in telehealth utilization across population groups may indicate that telehealth addresses some barriers to conventional health care while persistent inequities remain. Participants with diagnosed depression or anxiety and those with a history of cancer had higher odds of telehealth use than those without these conditions. These findings are consistent with AMA analyses showing substantial telehealth use in psychiatry; in 2024, 85.9% of psychiatrists reported having used video visits, and 56.9% reported that video visits accounted for more than 20% of their weekly visits^39^. These patterns suggest that telehealth may be particularly useful for patients with conditions requiring frequent or long-term care that can be delivered remotely. Telehealth coverage and reimbursement policies could therefore consider the needs of populations who may particularly benefit from continued access to remote care. However, the observed associations indicate only that participants with these conditions were more likely to use telehealth; the HINTS data do not establish whether telehealth was used specifically for the reported condition. Furthermore, higher utilization does not necessarily indicate greater clinical benefit. Future studies comparing clinical outcomes and quality of care between telehealth and in-person care could help determine when and for whom telehealth provides an effective alternative or complement to in-person services.

Additionally, the potential benefits of telehealth may not be equally accessible across populations. Adults aged 35 to 49 years had the highest adjusted odds of telehealth use, whereas similarly elevated use was not observed among the oldest adults. This pattern is notable because older adults generally have greater health care needs and may particularly benefit from reduced travel and more convenient follow-up care^40^. Telehealth utilization also increased with educational attainment, with the lowest use among adults with less than a high school education. These differences may reflect barriers identified in previous studies, including limited internet access, lower digital literacy, and greater difficulty navigating technology-enabled care, rather than the effects of age or educational attainment themselves^33,41^. Insured adults were also more likely to use telehealth than uninsured adults, suggesting that coverage and affordability may remain important determinants of access. Together, these patterns suggest that although telehealth can reduce some barriers associated with conventional in-person care, its use may also be constrained by barriers related to technology, digital readiness, and affordability^42^.

Geographic differences in telehealth utilization further suggest that access may not be uniform across the United States. Utilization was relatively low in the Midwest compared with other regions. Previous studies have documented geographic differences in telehealth access and identified digital infrastructure as one potential contributor11 ^43^. For example, rural areas have substantially less access to broadband infrastructure than urban areas, with fixed broadband availability reported to lag urban and suburban areas by 54%^17^. However, HINTS does (did?) not provide the state-level, rural–urban, broadband access, or device availability information needed to determine the mechanisms underlying the regional differences observed in this study. Therefore, lower utilization in the Midwest should not be interpreted as evidence of lower telehealth use among rural populations or attributed directly to differences in broadband access. Future studies incorporating state-level and rural–urban information together with measures of digital access could better identify the mechanisms underlying geographic variation in telehealth utilization. From a policy perspective, lower utilization in a region should not necessarily be interpreted as indicating lower need for telehealth. Telehealth policies and implementation strategies should consider whether limited digital infrastructure or technical access contributes to lower uptake and, where appropriate, support investments in digital infrastructure and technical assistance to ensure that populations in these regions have equitable opportunities to access telehealth services.

### Limitations

This study has several limitations. First, the available sample size limited our ability to examine telehealth modalities separately, including video, telephone, and combined video and telephone use, although these modalities may differ in technological requirements, accessibility, and clinical applications. Second, HINTS 6 and HINTS 7 are independent cross-sectional surveys rather than a longitudinal study; therefore, differences between 2022 and 2024 represent population-level patterns and should not be interpreted as changes in individual behavior. Third, HINTS did not collect comparable information on in-person health care utilization, limiting our ability to determine whether lower telehealth utilization reflected substitution by or return to in-person care. Fourth, the available data did not capture several factors that may influence telehealth use, including digital literacy, broadband and device access, provider availability, and state-level telehealth policies. Therefore, although differences in utilization were observed across sociodemographic and geographic groups, the mechanisms underlying these differences could not be directly evaluated. Despite these limitations, the use of nationally representative data, complex survey methods, and multiple imputation strengthened our assessment of telehealth utilization study.

## Conclusions

Telehealth utilization declined between 2022 and 2024, while the relative persistence of convenience as a reason for use suggests a shift away from pandemic-driven demand toward more selective integration into routine health care. Differences in utilization across population subgroups indicate that telehealth may address certain barriers to conventional care while access remains uneven. These findings highlight the evolving role of telehealth after the COVID-19 public health emergency and the need for policies that support its appropriate and equitable integration into routine care.

## Data Availability

All data produced are available online at The Health Information National Trends Survey (HINTS) website.

https://hints.cancer.gov/

## Author Contributions

Dr.Lin Ge had full access to all of the data in the study and takes responsibility for the integrity of the data and the accuracy of the data analysis.

Concept and design: All authors.

Acquisition, analysis, or interpretation of data: All authors.

Drafting of the manuscript: Wang.

Critical review of the manuscript for important intellectual content: Lin, Ge.

Statistical analysis: Wang.

Obtained funding: Ge.

Administrative, technical, or material support: Lin, Ge.

Supervision: Lin, Ge.

## Conflict of Interest Disclosures

The authors reported no relevant financial interests, activities, relationships, or affiliations related to the submitted work or during the 3 years before submission. No other relationships or activities that could be perceived to have influenced the submitted work were reported.

## Funding/Support

Publication of this article was supported by faculty start-up funds from School of Public Health-Bloomington, Indiana University.

## Role of the Funder/Sponsor

The funder had no role in the design and conduct of the study; collection, management, analysis, and interpretation of the data; preparation, review, or approval of the manuscript; and decision to submit the manuscript for publication.

## Data Sharing Statement

The HINTS 6 and HINTS 7 public-use data and documentation are publicly available from the National Cancer Institute Health Information National Trends Survey website.

## Notes

### Competing Interest Statement

The authors have declared no competing interest.

